# Impact of HIV and recreational drugs on cognitive functions in young men having sex with men

**DOI:** 10.1101/2022.04.25.22274123

**Authors:** Sophie Henrard, Nicola Trotta, Antonin Rovai, Tim Coolen, Hichem Slama, Julie Bertels, Delphine Puttaert, Jean-Christophe Goffard, Jean-Paul Van Vooren, Serge Goldman, Xavier De Tiège

**Author notes:** Corresponding author and reprint request: Dr Sophie Henrard, Department of Internal Medicine and immunodeficiency, Hôpital Universitaire de Bruxelles (H.U.B., site Erasme), Université libre de Bruxelles (ULB), Brussels, Belgium.

## Abstract

**Objectives:** This study characterizes the structural and metabolic cerebral correlates of human immunodeficiency virus (HIV)-associated neurocognitive disorders (HAND) in a preclinical setting that considers the lifestyle of young European men exposed to HIV, including recreational drugs.

**Design:** Prospective inclusion of participants.

**Methods:** Simultaneous structural brain magnetic resonance imaging (MRI) and positron emission tomography using [18F]-fluorodeoxyglucose (FDG-PET) were acquired on a hybrid PET-MRI system in 23 asymptomatic young men with HIV+ (mean age: 33.6 years, age range: 23-60 years; normal CD4+ cell count, undetectable viral load). Neuroimaging data were compared with that of a group of 26 young HIV-men, highly well matched for what concerns age, lifestyle, named pre-exposure prophylaxis users (HIV-PrEP), and to a group of 23 undifferentiated matched young men (i.e., healthy controls). A comprehensive neuropsychological assessment was also administered to the HIV+ and HIV-PrEP subjects.

**Results:** HIV+ subjects had lower performances in executive, attentional and working memory functions compared to HIV-PrEP subjects. No structural or metabolic differences were found between those two groups. Compared to healthy controls, HIV+ and HIV-PrEP exhibited a common frontal hypometabolism in the right prefrontal cortex that correlated with the level of recreational drug use. No structural brain abnormality was found.

**Conclusion:** A dynamic prevention of recreational drugs use in HIV+ and HIV-PrEP subjects is mandatory to cope with their negative impact on brain function and their neurocognitive consequences. A complex interplay between recreational drugs and HIV might be involved in the development of neurocognitive disorders in young men with HIV.

## Introduction

Even if the combined antiretroviral therapy (cART) dramatically reduced the incidence of dementia in people living with human immunodeficiency virus (HIV, *HIV+ subjects*), milder forms of cognitive impairments (i.e., HIV-associated neurocognitive disorders (HAND)) occur in 26-76% of HIV+ subjects (1).

HAND is mainly characterized by reduced attention, impairments in memory and executive functions (2). Executive functions corresponds to cognitive processes required to effectively initiate, monitor, and regulate thoughts and actions, thereby allowing one to respond in an adaptive manner to novel or complex situations (3). The dysexecutive syndrome associated with HAND is a predictor of employment status, risky sexual behavior, and cART adherence (4).

Neuroimaging techniques have been used to bring insights into HAND pathophysiology. Structural magnetic resonance imaging (MRI) revealed signs of cortico-subcortical atrophy in patients with HAND, even in those under cART (5). Few positron emission tomography (PET) studies using [18F]-fluorodeoxyglucose (FDG-PET) revealed reductions in mesial frontal/anterior cingulate cortex (ACC) (6, 7) or thalamic (8) glucose metabolism in asymptomatic HIV+ subjects under cART. HAND thus seems to be mainly associated with a dysfunction of the frontal lobe and its cortico-subcortical circuits. Still, such abnormalities may also be found in the context of recreational drugs use (RDU, (9), which is highly prevalent in a specific subset of HIV+ subjects during chemical sex (10), i.e., men having sex with men (MSM). RDU consumption therefore represents a major confound in studies addressing HAND pathophysiology (9).

This study aims at providing novel insights into HAND pathophysiology by combining clinical, neuropsychological, and lifestyle evaluations with hybrid PET and MRI (PET-MR) investigations in highly selected and matched populations of HIV+ and HIV-young men. To specifically disentangle the effects of HIV, cART and lifestyle (i.e., RDU) on cognitive, structural and metabolic brain changes, we first compared data of HIV+ subjects with those of a highly selected group of pre-exposure prophylaxis (PrEP) users (*HIV-PrEP users*) sharing similar lifestyle as MSM living with HIV. PrEP is a novel HIV prevention approach (11) relying on cART (tenofovir disoproxil (TDF), emtricitabine (FTC)). HIV-PrEP users represent an ideal group to disentangle HIV-related from lifestyle and cART effects on brain structure and function. We then compared data of HIV+ subjects and HIV-PrEP users with those of HIV-men taken from the general population (*healthy controls*) to highlight potential commonalities in cognitive/brain abnormalities between HIV+ subjects and HIV-PrEP users.

## Material and Methods

Methodological details can be found in Supplemental Digital Content (SDC). Only the essential information is provided hereunder.

### Participants

From December 2017 to June 2020, 25 western European, and native French-speaking HIV+ men (mean age: 38.8 years, age range: 18-60 years, >9 years of education) not declaring any cognitive impairment, were prospectively included. The exact duration of HIV infection was available for all of them (either the diagnosis was done during the acute infection or the presumed duration was based on negative HIV screening close to HIV diagnosis). All HIV+ subjects were taking cART with undetectable viral load for at least 6 months before inclusion and had a CD4+ cell count greater than 500/mm3. Those with a coinfection with hepatitis C, untreated syphilis, psychiatric disease, efavirenz based regimen (cART with brain side effects), or any history of brain disorder were excluded.

A total of 51 men matched for age with HIV+ subjects were also included. Those participants satisfied to the same criteria in terms of age, education and ethnicity as the HIV+ group. 26 of them were selected from the HIV-PrEP outpatient clinic. Fourteen HIV-PrEP users were taking TDF/FTC daily (30 pills/month), while 11 were taking it on demand before having a risky sexual intercourse (mean number of 15 pills/month) (11). The other 25 men were selected from the general population and constituted the healthy controls group based on their clinical background, lifestyle, and normal cognitive evaluation.

Participants contributed to the study approved by the institutional Ethics Committee after written informed consent (References: P2017/541, CCB: B4062017344197).

### Clinical and neuropsychological assessment

The clinical evaluation included an exhaustive anamnesis in all participants to collect their medical and psychiatric background. HIV+ subjects and HIV-PrEP users also underwent a comprehensive neuropsychological assessment investigating the main cognitive functions such as working memory, long-term episodic memory (on both verbal and visual material), gestural and visuo-constructive praxis, language (verbal fluencies and picture naming), attentional and executive functions. Questionnaires were used in all participants to assess depression, anxiety, previous night of sleep and cognitive functioning in daily living. Healthy controls’ broad cognitive functions were assessed with the Montreal cognitive assessment (MoCA) (12). RDU was evaluated in all participants through a semi-directed interview inspired by “the interview for research on addictive behavior” (13) that evaluated the frequency, the chronicity and the active/past character of the consumption of a series of drugs commonly used by MSM in the context of chemical sex (i.e., cocaine, synthetic and crystallized methamphetamine, ketamine, gamma-hydroxybutyrate/gamma-butyrolactoneheroin (GHB/GBL), poppers, cannabis) (14). An operational composite score (*RDU score*) was computed and incorporated the current and past exposition to recreative drugs. It was computed by adding the number of RDU on a monthly basis at the time of the study as well as those used on a monthly basis in the past (RDU= number of drugs used in the past + number of drugs currently used). This quantification was however a coarse-grained appreciation of the actual and past exposure as the majority of participants were unable to precisely evaluate the frequency and dosage of each current and past recreational drug due to the high variability in their use. Furthermore, to assess the potential toxicity of TDF/FTC (i.e., the cART shared by HIV+ subjects and HIV-PrEP users) on brain structure and function, an index was computed based on the dosage since the start of use (dosage=number of months of use). For HIV-PrEP users taking on demand regimen, the number of months of use was divided by two based on a mean number of fifteen pills/month (17).

### Analyses of neuropsychological data

To classify our participants according to Frascati’s criteria, individual performance of HIV+ subjects and HIV-PrEP users in the different tasks was then expressed as z-scores calculated by comparison with normative data. Here, we calculated z-scores for each task by subtracting the mean score of the normative data from a participant raw score, and then dividing the difference by the standard deviation of the normative data. For between-group comparison, we computed composite and index scores to quantify cognitive processes and reduce the number of data and variables. Scores of HIV+ subjects and HIV-PrEP users were compared using Student’s *t*-tests. Multiple comparisons were controlled using the Bonferroni correction. Relationships between cognitive scores and indexes, and TDF/FTC index or RDU score were assessed using Pearson’s correlations.

### PET-MR data acquisition

Cerebral FDG-PET and structural MRI data were obtained simultaneously using a 3T hybrid PET-MR scanner (SIGNATM, GE Healthcare). Structural MRI sequences for parenchymal evaluation (whole-brain axial 3D T1-weighted imaging (WI), axial T2WI, sagittal 3D T2 Fluid-Attenuated Inversion Recovery, axial 3D susceptibility-WI, and axial diffusion-WI) were acquired as previously described (15). Intracranial arteries were evaluated in HIV+ subjects to exclude active primary angiitis of CNS related to HIV status (16) by means of an axial 3D time-of-flight sequence, along with a pair of 3D T1-weighted “Black Blood” volumes, acquired before and after gadolinium-based contrast medium injection. For PET data acquisitions conducted as previously described (21), participants fasted for at least 4 hours, were awake in an eye-closed rest, and received an intravenous bolus injection of 3-5 mCi (111-185 MBq) of FDG. Forty minutes post-injection, a 20-min data acquisition was performed.

### MRI data preprocessing and analyses Qualitative analyses

MRI data were reviewed by one experienced neuroradiologist (T.C.) following a systematic and comprehensive visual assessment procedure searching for chronic cerebral small vessel disease and other parenchymal abnormalities, including extra-axial masses.

### Quantitative analyses

The Freesurfer 6 image analysis suite (http://surfer.nmr.mgh.harvard.edu/) was first used to obtain cortical volume and thickness using conventional surface-based approach (17-19) as well as to extract brain regions of interest (ROIs) based on anatomical brain parcellation (see supplemental table 1 (20, 21).

Student t-tests were used to search for differences in (i) maps of whole brain cortical volume and thickness and (ii) ROIs averaged volume (for cortical and subcortical ROIs) and thickness (for cortical ROIs only). These differences were tested between HIV+ subjects (taken individually or as a group) compared to HIV-PrEP users, and in HIV+ subjects and HIV-PrEP users (taken as separate or one group(s)) with that of healthy controls. For whole brain surface-based analyses, statistical maps were corrected for multiple comparisons using the False Discovery Rate (FDR) as implemented in Freesurfer 6, with the significance level set to p<0.05. For ROIs analyses, significance was considered at p<0.05 Bonferroni-corrected for the number of ROIs.

### PET data preprocessing and analyses

FDG-PET data were preprocessed and analyzed using the voxel-based Statistical Parametric Mapping software (SPM8, http://www.fil.ion.ucl.ac.uk/spm/, Wellcome Trust Centre for Neuroimaging, London, UK) based on conventional preprocessing, (individual and group level) subtractive and correlation analyses previously described (22-25).

Subtractive analyses identified brain areas where glucose metabolism was significantly lower or higher in HIV+ subjects (taken individually or as a group) compared with HIV-PrEP users. Additional group-level analyses compared the regional cerebral glucose metabolism of HIV+ subjects and HIV-PrEP users (taken as separate or one group(s)) with that of healthy controls. To search for a pathophysiological link between significant hypo- and hypermetabolic brain areas found in HIV+ subjects and HIV-PrEP users compared with healthy controls, we also performed pathophysiological interaction (*PPI*) analyses as previously described (25, 27). We finally performed correlations between the regional cerebral glucose metabolism of HIV+ subjects and HIV-PrEP users taken as one group and RDU score, TDF/FTC index, and neuropsychological data. Results were considered significant at p < .05 corrected for multiple comparisons over the entire brain volume (Family Wise Error (FWE)) or at a more liberal threshold (p < .001 uncorrected, height threshold: 0.001, cluster size k ≥ 50 voxels; or small-volume-corrected *P*<0.05 using a 10-mm radius spherical volume of interest).

## Results

### Clinical characteristics

Table 1 describes the clinical characteristics of each group of participants.

**Table 1.** Demographic and neuropsychological data.

|  | HIV+ (n=23) | PrEP (n=26) | Healthy controls (HC) (n=23) | p value |
| --- | --- | --- | --- | --- |
| Age (years) | 38 +/- 9.5 | 39 +/- 9.08 | 34 +/- 10.03 | HIV+/HC: p=0.17<br>HIV+/PrEP; p=0.75<br>PrEP/HC; p=0.08 |
| Duration of HIV infection (months) | 58 +/- 46.74 | / | / | / |
| CD4+ (cell count /mm3) | 834 +/- 294.83 | / | / | / |
| Timespan with CV>40 (rna copy/ml) | 31.11 +/- 41.69 | / | / | / |
| Recreative drug use (RDU) score | 4.21 +/- 3.26 | 4.78 +/- 3.51 | 0.54 +/- 0.59 | HIV+/HC: p<0.001<br>HIV+/PrEP; p=0.58<br>PrEP/HC; p<0.001 |
| TDF index (nb of pills) | 45.30 +/- 45.94 | 11.40 +/- 10.12 | / | P<0.001 |
| **Cognitive evaluation** |  |  |  |  |
| Nb of altered cognitive field | 1.70 +/- 1.02 | 1.44 +/- 0.87 | / | p=0.26 |
| HAND status |  |  |  |  |
| ANI | 21 | 23 | 0 | / |
| MND | 0 | 0 | 0 | / |
| HAD | 0 | 0 | 0 | / |
| MoCA | / | / | 28 +/- 1.27 | / |
| SRT | 12.177 +/- 1.261 | 12.360 +/- 0.9 | / | p=0.559 |
| Doors test | 18.565 +/- 2.446 | 19.654 +/- 2.465 | / | p=0.128 |
| Stroop test: Color IES | 60.384 +/- 10.931 | 66.283 +/- 54.085 | / | p=0.480 |
| Stroop test: Word IES | 43.083 +/- 5.816 | 46.273 +/- 33.824 | / | p=0.613 |
| Stroop test: Interference IES | 41.660 +/- 23.852 | 20.912 +/- 61.350 | / | p=0.038 |
| Stroop test: Flexibility IES | 137.430 +/- 36.52 | 115.120 +/- 20.975 | / | p=0.011 |
| TOL 2 IES | 75.035 +/- 97.387 | 31.285 +/- 23.604 | / | p=0.031 |
| TAP : AL_CV | 0.163 +/- 0.054 | 0.127 +/- 0.036 | / | p=0.01 |
| TAP : DS3 IES | 681.027 +/- 104.747 | 617.877 +/- 71.356 | / | p=0.019 |
| TAP : WM ME3 IES | 727.953 +/- 172.512 | 641.621 +/- 146.963 | / | p=0.009 |
| **cART** |  |  |  |  |
| Triumeq<br>Abacavir+lamivudine+dolutegravir | n=4 | / | / | / |
| Odefsey<br>Rilpivirine+emtricitabine+tenofovir (TAF) | n=9 | / | / | / |
| Biktarvy<br>Bictegravir+emtricitabine+tenofovir (TAF) | n=4 | / | / | / |
| Genvoya<br>Elvitegravir+cobicistat+emtricitabine+tenofovir (TAF) | n=3 | / | / | / |
| Descovy+Tivicay<br>Emtricitabine+tenofovir+dolutegravir | n=4 | / | / | / |
| Rezolsta+Tivicay<br>Darunavir+cobicistat+dolutegravir | n=1 | / | / | / |

No significant difference was found between HIV+ subjects and HIV-PrEP users in age, levels of anxiety/depression/quality of sleep, RDU score, and mean number of abnormal neuropsychological fields. Based on Frascati’s criteria, only 2 HIV+ subjects and 2 HIV-PrEP users were classified as normal, while 21 HIV+ subjects and 23 HIV-PrEP users were classified as having asymptomatic neurocognitive impairment (ANI) as they did not report any cognitive complain but were impaired in 2 or more cognitive domains (>1.0 standard deviation (SD) below the mean in a demographically appropriate normative mean). At the group level, performance was significantly lower in HIV+ subjects compared with HIV-PrEP users in executive functions (inhibition, p_corr_=0.038; combined EF, p_corr_=0.011; planification, p_corr_=0.031), attentional functions (vigilance, p_corr_=0.01; divided attention, pFWE=0.019) and working memory (WM updating, p_corr_=0.009), see Fig. 1. No correlation was found between RDU score and neuropsychological performance. Significant correlations, were found between TDF/FTC index and executive functions (combined EF, p_corr_=0.005, r=0,40; planification, p_corr_=0.039, r=0,3) in the form of a negative impact of TDF/FTC on executive functions.

**Figure 1.**
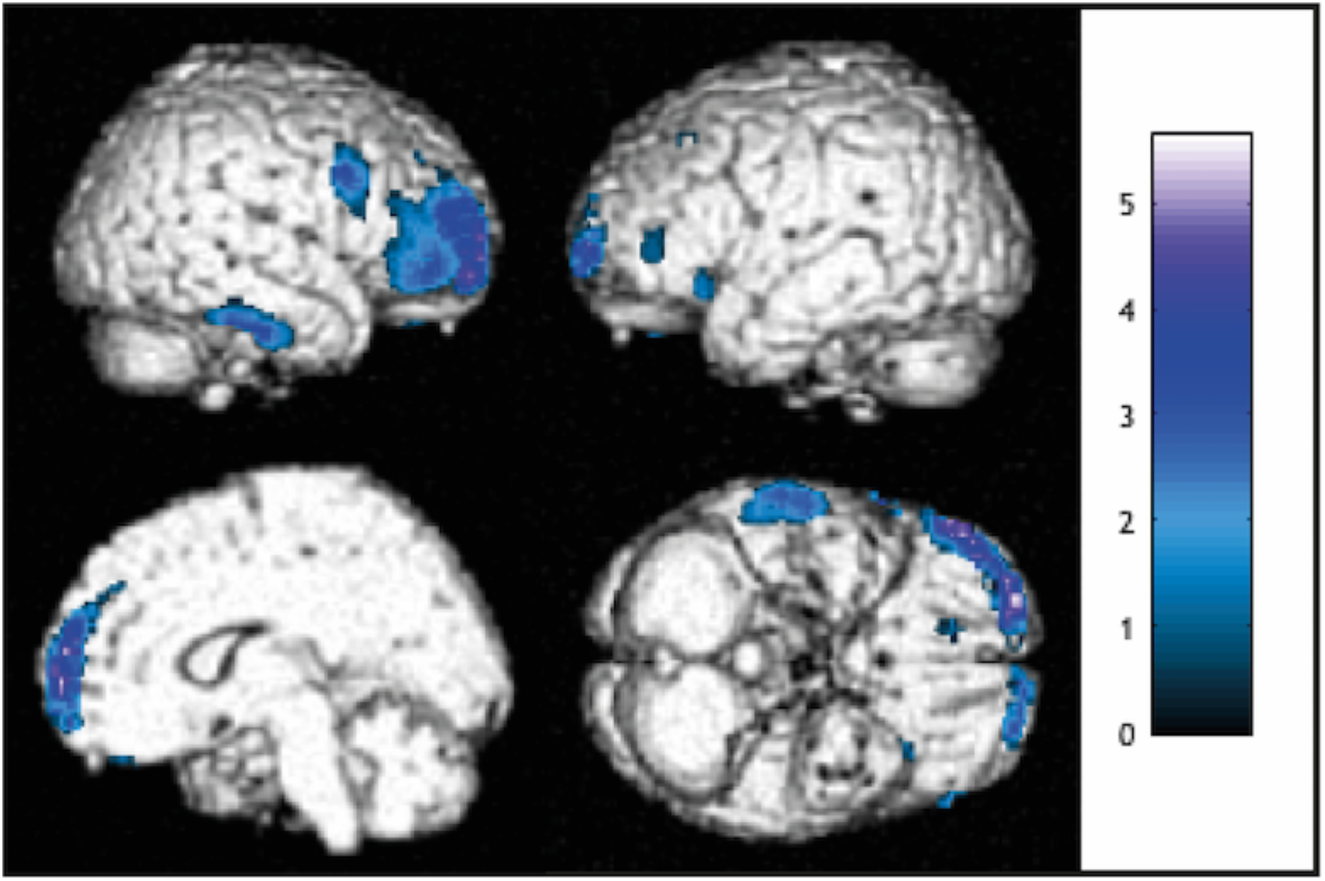
Subtractive analyses comparing FDG-PET data of HIV+ subjects and HIV-PrEP users taken as a group with those of healthy controls. Significant relative hypometabolic areas were observed in dorso-lateral prefrontal cortex bilaterally, the right dorso-medial prefrontal cortex, as well as in the left prefrontal and insular cortices.

### Structural MRI analyses

#### Qualitative analysis

Two HIV+ subjects, 1 HIV-PrEP user and 2 controls were excluded from further analyses due to incidental brain pathology or incomplete PET-MR acquisition due to a panic attack for one subject. The final sample was therefore composed of 23 HIV+ subjects, 25 HIV-PrEP users and 23 healthy controls.

#### Quantitative analysis

No significant difference (whole brain and ROI analysis) was found at the individual or group levels either with cortical volume, surface maps between HIV+ subjects and HIV-PrEP users, or between HIV+ subjects/HIV-PrEP users taken as one group and healthy controls.

### Regional cerebral glucose abnormalities

Figures 1-2 and Supplementary Tables 2-3 describe the group-level regional cerebral glucose abnormalities.

**Figure 2.**
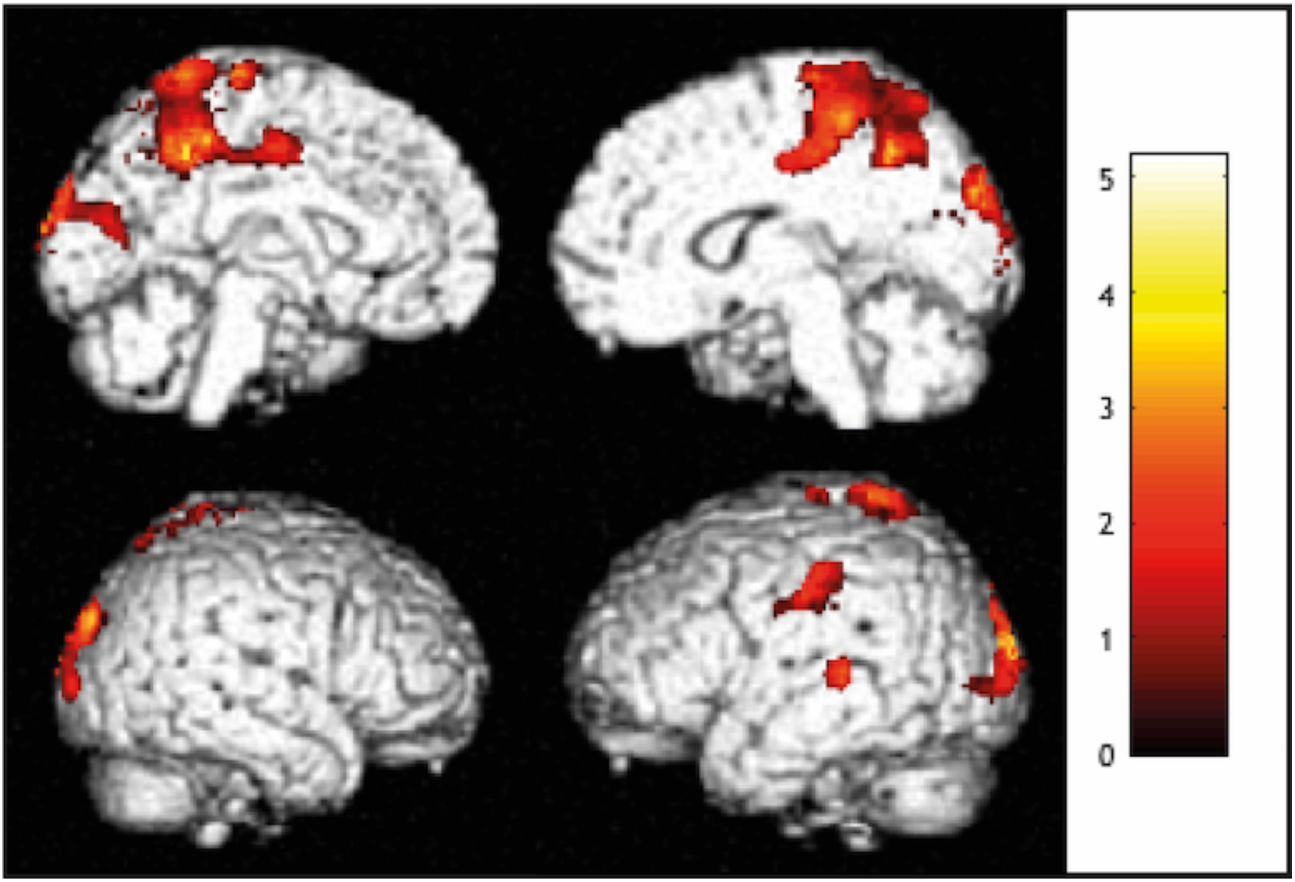
Subtractive analyses comparing FDG-PET data of HIV+ subjects and HIV-PrEP users taken as a group with those of healthy controls. Significant relative hypermetabolic areas were observed in the precuneus and cuneus bilaterally, the right primary somatosensory and posterior cingulate cortices, as well as the left superior temporal gyrus.

No group-level difference in regional cerebral glucose metabolism was found between HIV+ subjects and HIV-PrEP users (even at p_uncorrected_<0.001). When HIV+ subjects and HIV-PrEP users were pooled in one single group and compared with healthy controls, significant (p_FWE_<0.05) hypometabolism was found in the lateral and mesial (right > left) prefrontal cortex as well as significant (p_FWE_<0.05) hypermetabolism in posterior midline cortices bilaterally (precuneus and posterior cingulate cortex (PCC)). Similar prefrontral hypometabolism was found when comparing the brain metabolism of HIV+ subjects or HIV-PrEP users with that of healthy controls. When considering the peak voxel value in the hypometabolic right dorso-lateral prefrontal (DLPFC, MNI coordinates: [60, 14, 28]) and dorso-mesial prefrontal (DMPFC, [16, 64, 4]) cortices found in the pooled group of HIV+ subjects and HIV-PrEP users, no significant (event at p_uncorrected_<0.001) correlation was found between the level of metabolism in those brain regions and the rest of the brain, suggesting that prefrontal hypometabolism and posterior midline hypermetabolism were unrelated.

Correlation analyses were then performed between regional cerebral glucose metabolism and RDU score or TDF/FTC index in HIV+ subjects/HIV-PrEP users taken as one group. These analyses revealed a significant (p_FWE_ <0.05) negative correlation between RDU score and the level of metabolism in a right prefrontal cluster (right DLPFC, [60, 14, 28], (p_FWE_<0.01), r=-0.51; right DMPFC, [16, 64, 4], (p_FWE_<0.01), r=-0.52), see figure 3). No significant correlations were found between TDF/FTC index and prefrontal cortex metabolism. Finally, we did not find any significant correlation between regional cerebral glucose metabolism and the score of neuropsychological tests that significantly differed between HIV+ subjects or HIV-PrEP users. Figure 4 illustrates the significant hypometabolism observed at the individual level in 6 HIV+ subjects compared with HIV-PrEP users in the DLPFC or DMPFC (2 at p_FWE_<0.05, voxel level; 2 p_FWE_<0.05, cluster level; 2 at p_uncorrected_<0.001).

**Figure 3.**
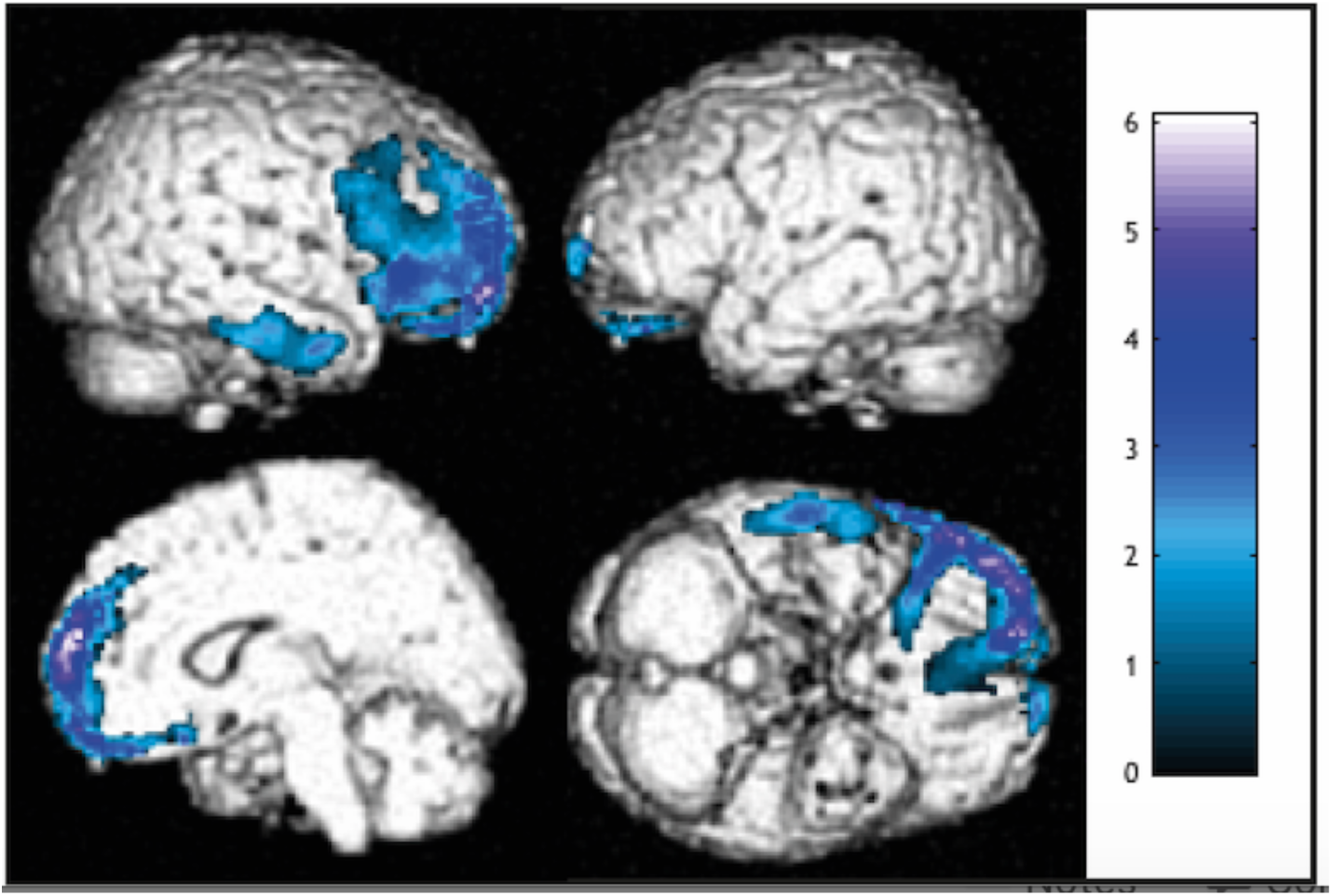
Results of the correlations analyses performed between the PET data of the HIV-PrEP and HIV+ and the cumulated drugs scores. Regression plots of cumulated drugs scores and adjusted metabolic responses were obtained by considering the peak voxel in (A) the right dorso-lateral prefrontal cortex [60, 14, 28] (Pearson’s correlation: r = -0.48 p < 0.001) and (B) the right dorso-medial prefrontal cortex [16, 64, 4] (Pearson’s correlation: r = -0.54, p < 0.001). These plots showed significant positive correlation between cumulated drugs scores and cerebral glucose metabolism in the considered voxels.

**Figure 4.**
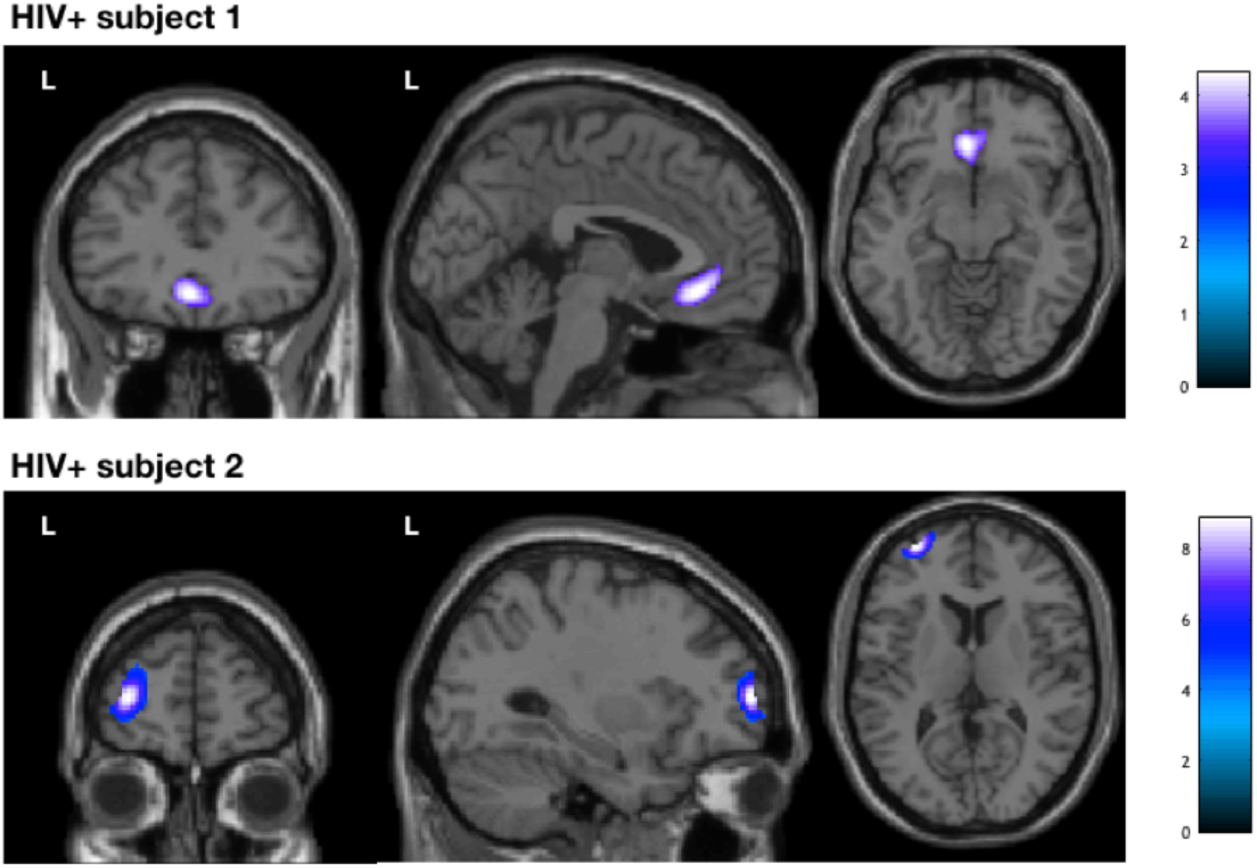
Subtractive analyses comparing FDG-PET data of HIV+ subjects taken individually with those of the group of HIV-PrEP users. Significant relative hypometabolic areas were observed in lateral or mesial prefrontal cortices of 25% of the HIV+. The figure shows for illustrative purposes two of these subjects exhibiting the aforementioned metabolic abnormalities.

## Discussion

Based on Frascati’s criteria, a high proportion (90%) of ANI was observed in HIV+ subjects and in HIV-PrEP users. Compared to HIV-PrEP users, HIV+ subjects also had significant alterations in executive, attentional and working memory functions in the absence of any group-level difference in brain structure and metabolism. Still, when each HIV+ subject was compared with HIV-PrEP users, 25% of them showed significant hypometabolism in prefrontal regions. Critically, when compared to healthy controls, HIV+ subjects and HIV-PrEP users displayed a significant prefrontal hypometabolism that correlated with their level of RDU but not with cognitive functioning.

### Neuropsychological assessment

Based on Frascati’s criteria, the prevalence of ANI (i.e., >1.0 to <2.0 SD below mean of a normative population in two or more domains but no decrease in everyday functioning) in our group of HIV+ subjects appeared especially high compared to the 26-76% previously reported (1). Surprisingly, the incidence of ANI was similar in our HIV-PrEP users. Thus, Frascati’s criteria do not optimally distinguish HIV+ subjects with HAND from HIV-subjects with similar life and health habits. They result in a high frequency of false-positive diagnosis due to the use of 1.0 SD deviation below mean, leading to an overestimation of HAND prevalence (1). Studies relying on those criteria might therefore suffer from an inadequate selection of neurocognitively impaired individuals. As previously suggested (1), our data further ask for the revisit of those criteria. The use of 2.0 SD below means of a normative population, as classically used in clinical neuropsychology, would be more appropriate to identify HIV+ subjects with HAND. Still, group-level analyses disclosed that HIV+ subjects had significantly lower performances in executive, working memory and attentional functions compared with HIV-PrEP users, which confirmed the dysexecutive/inattentive profile of HIV+ subjects previously reported (2).

### PET/MR results

Group-level analyses did not reveal any difference in regional brain structure or glucose metabolism between HIV+ subjects and HIV-PrEP users. This finding questions the pathophysiological role of HIV in the prefrontal hypometabolism found in HIV+ subjects when compared with healthy controls selected from the general population. Still, individual-level analyses showed that 25% of HIV+ subjects exhibited lateral or mesial prefrontal hypometabolism when compared with HIV-PrEP users. These changes at the individual-level were in line with the localization of the relative hypometabolism previously described in HIV+ subjects (6-9). The prefrontal hypometabolism found in some HIV+ subjects might potentially relate to the abnormalities noted in executive functions, although prefrontal metabolism did not significantly correlate with cognitive scores. Alternatively (but not exclusively), this hypometabolism, especially in the ACC, might also play a causative role in the adoption of high-risk behaviors in a drug and sexually-transmitted disease setting. Indeed, this brain area plays a pivotal role in decision making and risks/benefits balancing when emotional drive and objective reasoning come into conflict (26, 27). It may thus be causative rather than the consequence of the contracted disease (26, 27). This hypothesis warrants further investigations. The absence of group-level metabolic difference between HIV+ subjects and HIV-PrEP users suggests that their common brain hypometabolic abnormalities in comparison to healthy controls might be driven by another cause than HIV. Correlation analyses pointed to the well-known vulnerability of the prefrontal areas to the effects of RDU (9, 28). In the absence of noticeable structural brain changes, hypometabolism was probably related to neural dysfunction rather than neuronal loss *per se*. RDU-induced neural dysfunction might be related to dopamine dysregulation, neuroinflammation or neurodegeneration through the accumulation of intraneuronal hyperphosphorylated TAU protein (pTAU) (29-31). In HIV+ subjects, recreational drugs might exert their neural toxicity synergistically with HIV-related neuroinflammation, neurodegeneration or the reactivation of deep latency virus (30, 32). A complex interplay between RDU and HIV might thus be involved in the induction and development of the prefrontal hypometabolism and dysexecutive/inattentive profile found in our HIV+ subjects. Still, results obtained in HIV-PrEP users suggest that RDU might play a predominant pathophysiological role compared with HIV. The absence of correlation between regional brain metabolism and cognitive alterations observed in our subjects might be explained by the inability of FDG to explore neuroinflammation or cerebral pTAU deposition. Other radioligands targeting neuroinflammation-related or pTAU proteins could thus be used and correlate with cognitive dysfunctions (33).

Hypermetabolism in posterior midline cortices (precuneus and PCC) belonging to the default mode network (DMN, (34) were also found at the group level in HIV+ and HIV-PrEP compared to healthy controls. Correlation analyses failed to find any relationship between the level of prefrontal hypometabolism and posterior midline cortices hypermetabolism, suggesting that these metabolic abnormalities have no pathophysiological link. The posterior midline cortices hypermetabolism, which did not correlate with RDU score, could be related to attention/dysexecutive alterations (35).

## Structural Results

At the preclinical stage of HAND, noticeable brain atrophy is not expected. Still, our data are in relative contradiction with the literature, which supports that, even at the preclinical stage, atrophy is detectable in caudate nuclei and prefrontal cortex of patients with HAND. This discrepancy may be related to the high heterogeneity of HIV+ subjects included in previous studies, especially regarding the mix of age, education level, comorbidities, viral loads and cognitive symptoms (13). Further studies should therefore concentrate on specific populations of HIV+ subjects, as done in the present study.

## Clinical Implications

Our findings strongly support the effect of lifestyle in MSM population and the importance of better screening/control strategies to limit the occurrence of neurocognitive disorders. Development and use of an international recreational drug scores, such as the multi-morbidity index (CCI) (36), might be of great clinical help to assess who is at higher risk of HAND. Our data also reinforces the need for future studies to include well selected HIV populations (exclusion of comorbidities and recreational drugs use) and proper control populations to properly address virus-driven pathogenesis in HAND in an undetectable virus setting. Our study also describes the brain metabolic abnormalities in HIV-PrEP users, highlighting their vulnerability for the development of neurocognitive changes to RDU. Still, the correlation observed between TDF/FTC index and executive functions suggests that these antiviral drugs, that are common to both HIV+ subjects and HIV-PrEP users, might contribute, in addition with RDU, to the development of ANI. As it is in contradiction with previously published data ((37, 38), this finding call for future investigations to clarify the pathophysiological role of these antiviral drugs in HAND.

## Limitations

The limited number of subjects included in this study, due to the strict inclusion criteria, may have impacted the statistical power of group-level subtractive and correlation FDG-PET analyses. The very homogeneous profile of our population is our main asset, but it inherently results in a limitation, i.e., our results might be inapplicable in another settings (e.g., African older comorbid women) since only Caucasian young men were investigated. Finally, we did not apply the same cognitive assessment to healthy controls than to the HIV+ subjects and HIV-PrEP users.

## Conclusion

This study disclosed a dysexecutive/inattentive profile and prefrontal hypometabolism in HIV+ subjects in the absence of noticeable brain atrophy. Prefrontal hypometabolism was similar to that observed in HIV-PrEP users and was related to RDU. A dynamic prevention of RDU in those populations is therefore warranted to cope with their negative impact on brain function and their neurocognitive consequences. A complex interplay between recreational drugs, the use of TDF/FTC, and HIV might be involved in the development of neurocognitive disorders in MSM with HIV.

## Data Availability

All data produced in the present study are available upon reasonable request to the authors

## Fundings and aknowledgments

Sophie Henrard and Delphine Puttaert have been supported by the Fonds Erasme (Brussels, Belgium). Tim Coolen has been a Clinical Master Specialist applicant to a PhD at the Fonds de la Recherche Scientifique (FRS-FNRS, Brussels, Belgium). Xavier De Tiège is Clinical Researcher at the FRS-FNRS.

The PET-MR project at the ULB has been supported by the Association Vinçotte Nuclear (Brussels, Belgium).

| Name | Location | Contribution |
| --- | --- | --- |
| Sophie Henrard, MD | Department of Internal Medicine, HIV Reference Center, HUB Erasme, Université libre de Bruxelles (ULB), Brussels, Belgium | Study concept and design; data acquisition and analysis; interpretation of data; full access to all of the data in the study; responsible for the |
|  |  | integrity of the data and the accuracy of the data analysis; drafting the manuscript; critical revision of the manuscript for important intellectual content. First authorship. Obtained funding. |
| Nicola Trotta, PhD | Laboratoire de Cartographie fonctionnelle du Cerveau, UNI —<br>ULB Neuroscience Institute, Université libre de Bruxelles (ULB), Brussels, Belgium; Department of Nuclear Medicine, CUB Hôpital Erasme, | Analysis and interpretation of data; critical revision of the manuscript for important intellectual content |
| Antonin Rovai, PhD | Laboratoire de Cartographie fonctionnelle du Cerveau, UNI —<br>ULB Neuroscience Institute, Université libre de Bruxelles (ULB), Brussels, Belgium; Department of Nuclear Medicine, CUB Hôpital Erasme, | Analysis and interpretation of data; critical revision of the manuscript for important intellectual content |
| Tim Coolen, MD | Department of Radiology, CUB Hôpital Erasme, Université libre de Bruxelles (ULB), Brussels, Belgium; Laboratoire de Cartographie fonctionnelle du Cerveau, UNI —<br>ULB Neuroscience Institute, Université libre de Bruxelles (ULB), Brussels, Belgium | Interpretation of data; critical revision of the manuscript for important intellectual content |
| Hichem Slama, PhD | Head of neuropsychology department and speech, HUB Erasme, Université libre de Bruxelles (ULB), Brussels, Belgium. | Neurocognitive data acquisition. Analysis and interpretation of data; critical revision of the manuscript for important intellectual content |
| Julie Bertels, PhD | ULBabyLab-Consciousness, Cognition and Computation Group, Center for Research in Cognition and Neurosciences, ULB Neuroscience Institute, Université libre de Bruxelles (ULB), Brussels, Belgium. | Neurocognitive data acquisition, critical revision of the manuscript for important intellectual content. |
| Delphine Puttaert, PhD | Laboratoire de Cartographie fonctionnelle du Cerveau, | Neurocognitive data acquisition, critical revision of |
|  | UNI — ULB Neuroscience Institute, Université libre de Bruxelles (ULB), Brussels, Belgium | the manuscript for important intellectual content. |
| Jean-Christophe Goffard MD, PhD | Head of department of Internal Medicine, HIV Reference Center, HUB Erasme, Université libre de Bruxelles (ULB), Brussels, Belgium | Study concept and design; critical revision of the manuscript for important intellectual content. |
| Jean-Paul Van Vooren, MD, PhD | Department of Internal Medicine, HIV Reference Center, HUB Erasme, Université libre de Bruxelles (ULB), Brussels, Belgium | Critical revision of the manuscript for important intellectual content. |
| Serge Goldman, MD, PhD | Laboratoire de Cartographie fonctionnelle du Cerveau, UNI — ULB Neuroscience Institute, Université libre de Bruxelles (ULB), Brussels, Belgium; Department of Nuclear Medicine, CUB Hôpital Erasme. | Study concept and design; interpretation of data; full access to all of the data in the study; responsible for the integrity of the data and the accuracy of the data analysis; drafting the manuscript; critical revision of the manuscript for important intellectual content. |
| Xavier De Tiège, MD, PhD | Laboratoire de Cartographie fonctionnelle du Cerveau, UNI — ULB Neuroscience Institute, Université libre de Bruxelles (ULB), Brussels, Belgium; Department of Nuclear Medicine, CUB Hôpital Erasme. | Study concept and design; interpretation of data; full access to all of the data in the study; responsible for the integrity of the data and the accuracy of the data analysis; drafting the manuscript; critical revision of the manuscript for important intellectual content. |

## Data availability statement

Anonymized neuropsychological and neuroimaging data can be obtained upon reasonable request to the corresponding author and after approval of institutional (Hôpital Universitaire de Bruxelles and Université libre de Bruxelles) authorities.

