## Supplementary material for "Impact of HIV and recreational drugs on cognitive functions in young men having sex with men": Supplemental .pdf

### Supplemental Digital Content

### Supplemental Methods

The Supplemental Methods aim at providing additional information and details about the methods used for the neuropsychological assessment and for neuroimaging acquisition and analyses.

### 1. Neuropsychological assessment

#### 1.1. Description of cognitive scores and indexes

To quantify cognitive processes and reduce the number of data and variables, we computed, depending on the tasks (see 1.2. for a complete description of the tasks), composite scores and index scores. Z-scores were computed for each cognitive task to classify participants (i.e., HIV+ subjects and HIV-PrEP users) relative to normative data according to the Frascati's criteria for HAND.

1.1. 1. Composite scores: Inverse Efficiency Scores

To integrate speed and accuracy, we chose to compute a composite score called the Inverse Efficiency Score (IES)(1, 2). The IES is a well-established index of integrated Speed-Accuracy measures. The IES is computed as the ratio of the mean or the median reaction time (RT) by the proportion of correct responses (PC). PC is also calculated as 1 minus the proportion of errors (PE):

$$IES = RT / PC = RT / (1-PE).$$

For instance, a patient completing a task in 60 seconds with an accuracy of 80% will have an IES of: 60 s / 0.8 = 60 s / (1 - 0.2) = 75 s. We decided to use median reaction times because they are less affected by extreme values than mean reaction times. A higher IES indicates a lower cognitive capacity.

1.1.2. Index scores: difference scores

Complex higher-order cognitive functions, such as working memory, attentional processes, and executive functions (EF), are difficult to assess using task performance because they exert their action through lower-level functions. Complex tasks are thus multidetermined and involve several cognitive functions and processes, which is coined in the literature as the task-impurity problem (3, 4). To estimate higher-order functions, neuropsychologists use simpler control tasks that assess whether basic cognitive processes are responsible or not for a deficit in a more complex task. To extract higher-order cognitive components from task performance, difference scores or ratio-scores are usually computed (5). Difference scores are current gold standards to assess inhibition (see for instance (6) for a review on the Stroop task) or cognitive flexibility (see (7, 8) for a review on task switching). The rational for computing difference scores is to subtract time or accuracy (or integrated scores such as IES) related to low-level cognitive processes from the performance in the complex higher-order task. Therefore, difference scores allow to isolate higher-order cognitive components. Here, we used the same rational to compute index scores for each complex task including a simpler control condition. A higher difference score indicates a lower cognitive capacity.

Performance in the complex task = Low-level cognitive processes + high-level cognitive processes

Performance in the complex task - Low-level cognitive processes = High-level cognitive processes

An alternative to a difference score is a ratio score between the complex and the simpler task. The major advantage of ratio scores is that they take individual differences in baseline processing speed (or accuracy) into account. Therefore, they reveal only abnormalities that exceed the contribution of general slowing. This can be viewed as a limitation that may not always be desirable. For instance, in a between-groups comparison, if general slowing does not affect the higher-order component (i.e., groups have comparable response speed in the control condition), the “true” alteration level may be underestimated when using ratio scores (5). Because our HIV population is not characterized by severe alterations and slowing, we decided to compute difference scores (Δ scores), not ratio scores.

#### 1.1.3. Calculating z-scores

The z-score or standard score is the number of standard deviations by which the value of a raw score (i.e., an observed value) is above or below the mean value of what is being observed or measured (9). Raw scores above the mean have positive standard scores, while those below the mean have negative standard scores. Here, we calculated z-scores for each task by subtracting the mean score of the normative data from a patient raw score, and then dividing the difference by the standard deviation of the normative data.

### 1.2. Cognitive functions, tests and questionnaires

#### 1.2.1. Working memory

The three components of Baddeley and Hitch's working memory (WM) model (10) were assessed: the phonological loop, the visuospatial sketchpad and the central executive. The phonological loop (verbal short-term memory, VSTM) was evaluated using a forward digit span (11). In this task, participants were asked to repeat sequences of digits (verbally presented and increasing in length) in the same order as during presentation. Digits were presented at the pace of one digit per second. There were eight difficulty levels (from two to nine digits), each comprising three sequences of digits. The task was interrupted after the three trials of a span level were failed. The total number of correct trials was used as the variable of interest because it is a more sensitive score than the highest span (12, 13). The visuospatial sketchpad (spatial short-term memory, SSTM) was evaluated with the Block-Tapping Test (14). In this task, participants were presented with nine blocks arranged on a board. The examiner tapped on several blocks and the participant had to tap on the same blocks, in the same order. The sequence started with two blocks and gradually increased up to nine blocks. We assessed two functions of the central executive of WM, i.e., manipulation of information in WM and information updating (15, 16). Manipulation of information was assessed using a backward digit span (11) where the sequences had to be repeated in the reverse order (manipulation of information in WM, WM manipulation). To estimate the central executive component, we computed a difference score between backward and forward digit span (ΔWM).

$$\Delta\text{WM manipulation} = \text{digit span forward} - \text{digit span backward}$$

Updating processes in WM were assessed using the 2-back task of the Test of Attentional Performance (TAP) computerized battery (17). Participant had to press a button when the digit currently displayed matched the digit presented two steps earlier in a visual sequence. Therefore, for successful comparison, the task required for successful comparison both the maintenance and the updating of a series of items in WM. An IES was computed for WM updating using the median RT and proportion of both omissions and errors.

$$\text{IES WM updating} = \text{median RT} / [1 - (\%\text{ errors} + \%\text{ omissions})/2]$$

1.2.2. Long-term episodic memory

Long-term episodic memory was assessed for verbal and visual modalities. Verbal episodic memory (VLTM) was assessed using a French adaptation of the Selective Reminding Test (SRT) (18, 19), and pictorial episodic memory (PLTM) was evaluated with the Doors and People test (visual recognition part) (20). The SRT is a test designed to measure verbal learning and episodic memory using a list-learning procedure over multiple trial. This paradigm is believed to separate verbal memory into distinct processes. The SRT involves reading the participant a list of 15 unrelated words and then having him immediately recall as many of these 15 words as possible. Every trial after the first involves selectively presenting only those words in which the participant did not recall on the immediately preceding trial. The selective reminding trials proceed in this manner until the participant can correctly recall all 15 words on two consecutive trials, or until 10 trials have been completed. Through assessing the recall of items that are not presented on a given trial, this test is believed to distinguish between retrieval from long-term memory (LTM) and short-term memory (STM). Words not recalled on two consecutive trials are believed to be recalled from STM. Words that have been recalled on two consecutive trials, but presented only on the first of the two trials, are believed to be recalled from LTM. A mean total recall score is calculated, which assesses the sum of STM and LTM. The words recalled from LTM are further broken down into those recalled on every subsequent trial (consistent long-term retrieval, CLTR) and those recalled inconsistently. A delayed recall score is also calculated after 30 minutes. We computed a Δ score to estimate consolidation processes.

| |
|---|
| VLTM score = mean of recall scores |
| VLTM CLTR = proportion of words recalled on every subsequent trial |
| VLTM delayed recall = number of words recalled after 30 minutes |
| ΔVLTM consolidation = last recall – delayed recall |

In the Doors and People test (visual recognition subtest) the participant attempts to memorize a series of 2 x 12 colored photographs of doors (parts A and B). Pictural episodic memory (PLTM) is tested by recognition of each target door from a set of four doors varying in similarity, and hence difficulty (part A is simpler than part B). The total score is the sum of part A and B.

| |
|---|
| PLTM score = sum of recognition scores fort parts A and B |

1.2.3. Executive functions

We assessed 3 core executive functions (EF) (WM described before, inhibition and cognitive flexibility) and their combination, and 2 higher-level EF (planning and reasoning) (21).

1.2.3.1. Inhibition

We used the Stroop test (22, 23) to assess inhibition processes. The color and word Stroop test was composed of 3 subtests of 100 items: color naming, color-word reading and interference. In the color naming task, participant was asked to name the color of rectangles. In the color-word reading task, participant was asked to read the color word. In the interference part, participant was asked to name the color of the ink in which each word was written. Because written words are incongruent color words, an interference effect occurs. For instance, the word "RED" is displayed in green. Usually, participant requires more time and are less accurate in responding "green" than in the color naming part.

We computed IES for the 3 parts of the Stroop test. After calculating IES for each subtest, we computed difference scores to estimate inhibition processes.

| Δ Inhibition = IES color-word condition – IES color condition. |
|---|

1.2.3.2. Cognitive flexibility

Cognitive flexibility was assessed using the Flexibility subtest of the TAP battery (17). In the Flexibility subtest of the TAP, one letter and one number appear side-by-side on the screen and switch positions randomly across trials. In the control parts, participant was told to indicate on which side of the screen the target figure (the letter in part 1 and the digit in part 2) appeared on by pressing either a left or right key on a button box. In the switching part (part 3), participant was told to alternate their focus between each of the items —first the letter, then the number, then the letter, and so on. We first computed an IES based on the number of errors and median reaction time (RT) for each part. Then, we calculated a flexibility index (i.e., a switch cost) by subtracting the mean IES for the two control parts from the IES for the flexibility part.

| Δ Flexibility = IES flexibility part – (IES letter part – IES digit part)/2 |
|---|

1.2.3.3. Combined EF

Core EF (WM, inhibition, flexibility) are both separable and correlated (21, 24). Therefore, we also used a complex executive task involving the 3 core EF at the same time to assess their combination:

the Stroop flexibility task. In the Stroop flexibility task, participant was asked to name the color of the ink in which each word was displayed, but if the word were framed, he had to read the color-word. The Stroop flexibility task thus involves inhibition because participant has to inhibit the incongruent response, flexibility because he has to switch between naming the ink color and reading the color word, and WM because they have to maintain the instruction during the task. We computed a Core EF IES and a Δ Core EF:

Core EF IES = Stroop flexibility RT / Stroop flexibility PC

Δ Core EF = Core EF IES – (color-word IES + color reading IES)/2.

1.2.3.4. Planning

Participant was administered the Tower of London (ToL) (25, 26) to assess planning abilities (higher-order EF) (21). The ToL consists of two boards with pegs and several beads with different colors. The neuropsychologist uses the beads and the boards to present the examinee with problem-solving tasks. Participant was asked to achieve the problem-solving task with the less moves they can. This task included three test trials.

A Planning IES for the ToL was computed to assess planning abilities. The subsequent time (time between the first and the last movement of a task item) was divided by the accuracy. Accuracy was computed as the ratio between the expected number (N) of moves and the number of moves made by the participant:

Planning IES = Subsequent time / [ (expected N of moves) / participant N of moves ].

1.2.3.5. Reasoning

Reasoning, a higher-order EF similar to the concept of fluid intelligence (21) was assessed with the Matrix subtest of the Wechsler Adult Intelligence Scale fourth version (WAIS-IV) (11). Participant viewed an array of pictures with one missing square and had to select the picture that fits the array from five options. The total score of the Matrix subtest was used for the analyses.

#### 1.2.4. Attentional functions

The computerized Test for Attentional Performance (TAP) battery (17) was used to assess attentional functions. Attentional abilities were assessed using the Alertness and Divided Attention subtests of the TAP. The alertness subtest is composed of two conditions: a simple reaction time task (tonic alertness) in which a cross appeared on the screen at random intervals and the participant had to respond as fast as possible by pressing a response button; and an auditory-cued reaction time task (phasic alertness) in which a warning tone preceded the apparition of the cross. In the divided attention subtest, the participant had to process simultaneously visual and auditory stimuli. The participant had to respond as fast as possible by pressing a response button when four crosses were forming a square (part 1; visual selective attention), when he heard two identical tones consecutively (part 2; auditory selective attention), and for both situation at the same time (part 3; divided attention).

To quantify tonic alertness, we used the median reaction time (RT) of the TAP alertness subtest (condition without auditory cuing) which is less affected by extreme RT. For vigilance, we computed the coefficient of variation of RT from the TAP alertness (condition without auditory cuing) that is the ratio of the RT standard deviation (SD) divided by the mean RT:

Tonic alertness index = median RT

Vigilance index = coefficient of variation = standard deviation of RT / mean RT

For divided attention, we computed IES for the three parts of the TAP subtest (visual selective attention, auditory selective attention, and divided attention). Then, we computed a Δ score to quantify divided attention.

$$\Delta \text{ Divided attention} = \text{IES divided attention} - (\text{IES visual selective attention} - \text{IES auditory selective attention}) / 2$$

1.2.5. Language

Language was assessed using verbal fluencies and object naming tasks (23, 27).

Verbal fluencies tasks are designed to assess the ability to retrieve words from the spelling lexicon (letter fluency) and from the semantic lexicon (semantic fluency). In the letter fluency task, participant was asked to retrieve as many words as he could that started with the letter P within 2 minutes. In the semantic fluency task, he has to retrieve as many animals as possible within 2 minutes. Spelling and semantic fluencies scores are the total number of words generated for each category.

Object naming abilities were assessed with the Lexis test (28). Participant had to name 80 drawings of objects. The total score (maximum = 80) was used for analyses.

1.2.6. Praxis functions

Visuo-constructive praxis were assessed with the Rey Complex Figure (RCF) (29). In the RCF, participants were asked to copy an abstract complex figure. The copy was scored for the copy accuracy and location of 18 elements of the figure. The total score (maximum score = 36) was used for analyses.

Gestural praxis was assessed with the Brief Screening Scale of Gestural Praxis (30). Participant had to execute 5 symbolic gestures on verbal command, to execute 5 action gestures on verbal command, and to reproduce 8 abstract gestures made by the neuropsychologist. The total score of the task (maximum = 23) was used for analyses.

1.2.7. Everyday life cognition

Everyday life cognition was assessed using the Cognitive Difficulties Scale (CDS) (31). The CDS is a 39 statements self-report questionnaire that asks participants to describe everyday inefficiencies, lapses of attention or memory and related functions that people notice about themselves. The questionnaire contains statements describing their typical or usual behavior; subject rate how often they have experienced these during the previous month on a five-point Likert scale (0 = never to 4 = very often); high scores indicate frequent and severe cognitive difficulties. Th total score (maximum = 156) was used for analyses.

1.2.8. Anxiety

Anxiety was assessed using the State-Trait Anxiety Inventory (STAI), a commonly used measure of trait and state anxiety (32). We used the Anxiety Form Y that has 20 items for assessing trait anxiety (part B) and 20 for state anxiety (part A). All items are rated on a 4-point scale (e.g., from “Almost Never” to “Almost Always”). Higher scores indicate greater anxiety.

1.2.9. Depression

Depression was assessed using the 13 items Beck Depression Inventory (BDI) (33). The BDI is a multiple-choice self-report inventory measuring the severity of depression.

1.2.10. Sleep

We assessed the quality of participant’s previous night of sleep with an adaptation of The St. Mary's Hospital sleep questionnaire (34).

1.2.12. Recreational drug use (RDU) score

RDU score was computed by adding the number of RDU on a monthly basis at the time of the study as well as those used on a monthly basis in the past (RDU= number of drugs used in the past + number of drugs currently used). This quantification was however a coarse-grained approximation of the actual and past exposure as the majority of participants were unable to precisely evaluate the frequency and dosage of each current and past recreational drug due to the high variability in their use.

1.2.12. Procedure

Tasks and questionnaires were administered by three experienced neuropsychologists (J.B.: 9 HIV+ subjects, 11 HIV-PreP users; D.P.; 11 HIV+ subjects, 11 HIV-PreP users; H.S.: 5 HIV+ subjects, 4 HIV-PreP users) in a single session of three hours. The neuropsychologists were blind to the HIV status of the participants. Tasks and questionnaires were administered in the following fixed order:

- Cognitive Difficulties Scale

- St. Mary's Hospital sleep questionnaire

- State-Trait Anxiety Inventory Part A (state anxiety)

- Selective Reminding Test

- Doors and People test (visual recognition)

- Stroop test

- Forward digit span

- Backward digit span

- Block-tapping test

- TAP working memory subtest

- TAP divided attention subtest

- TAP alertness subtest

- TAP flexibility subtest

- Tower of London

- Letter and semantic fluencies

- Lexis test

- Brief Screening Scale of Gestural Praxis

- Rey Complex Figure test

- WAIS-IV Matrix subtest

- State-Trait Anxiety Inventory Part B (trait anxiety)

- Beck Depression Inventory

### 2. Neuroimaging data acquisition, preprocessing and analyses

#### 2.1. PET-MR data acquisition

Cerebral FDG-PET and structural MRI data were obtained simultaneously using a 3T hybrid PET-MR scanner (SIGNA™, GE Healthcare, Chicago, IL, USA) as in previous publications (35, 36).

MRI sequences consisted of whole-brain axial 3D T1-weighted imaging (WI, repetition time (TR)/echo time (TE)/Flip Angle (FA): 8ms/3ms/12°, inversion time (TI): 450ms, field of view (FOV): 24cm x 24cm, matrix: 240 x 240, resolution: 1mm x 1mm x 1mm), axial T2WI (TR/TE/FA: 6500ms/126ms/142°, FOV: 24cm x 24cm, matrix: 480 x 480, resolution: 0.5mm x 0.5mm x 3mm, slice spacing: 0.3mm), sagittal 3D T2WI FLAIR (TR/TE/FA: 7200ms/120ms/90°, TI: 1333 to 2041 ms, FOV: 25.6cm x 25.6cm, matrix: 256 x 256, resolution 1mm x 1mm x 1.4mm), axial 3D SWI (TR/TE/FA: 48ms/25ms/10°, FOV:24cm x 24cm, matrix: 240 x 240, resolution: 1mm x 1mm x 1.6mm), and axial DWI (TR/TE/FA: 6500ms/80ms/90°, FOV: 26cm x 26cm, matrix: 128 x 128, resolution: 2mm x 2mm x 4mm, slice spacing: 0.4mm). In order to exclude primary angiitis of the CNS (see exclusion criteria) in HIV+ subjects, we also acquired in these subjects a set of two axial 3D Black Blood images (Cube Vessel Wall with TR 650ms, TE min, ETL 30, BW 62.50kHz, 1mm isotropic voxels and Motion-Sensitized Driven-Equilibrium (MSDE) pulse set to suppress speed below 3cm/s in all directions) acquired with and without gadolinium (Dotarem) injection, as well as a 3D Time-Of-Flight (TOF with FA 15°, Out-of-phase TE, one echo, TR 30msn BW 41.67kHz, 1mm isotropic voxels) image.

For PET data acquisitions, participants fasted for at least 4 hours, were awake in an eye-closed rest and received an intravenous bolus injection of 3-5 mCi (111-185 MBq) of FDG before PET-MR data acquisition. Forty minutes post-injection, a 20-min data acquisition was performed. PET data were reconstructed using the fully 3D iterative reconstruction algorithm VUE Point FX-S, which considers the time-of-flight (TOF) information and the correction for the point spread function (PSF) of the system. The algorithm was configured with 10 iterations, 28 subsets and a standard Z-axis filter cut-off at 4 mm. The photons' attenuation was corrected with an MRI-based map (MRAC) acquired simultaneously. PET images were displayed in a 256 x 256 x 89 matrix format, with a slice thickness of 2.78 mm. The reconstructed files were downloaded in their original format (DICOM, ECAT, Interfile) for meta- information and converted in NIfTI format for subsequent analysis.

### 2.2. MRI data preprocessing and analyses

#### *2.2.1. Qualitative analyses of MRI data*

MRI data were reviewed by one experienced neuroradiologist (T.C.) following a systematic and comprehensive visual assessment procedure. First, lesions attributed to chronic cerebral small vessel disease were evaluated including white matter hyperintensities on FLAIR imaging graded according to the Fazekas scale (37), and the presence of lacunes on T2WI and FLAIR as well as microbleeds on the SWI sequence. Larger sequellar lesions were also searched for in the parenchyma and labeled postischemic unless they corresponded to a posttraumatic pattern (38) or were prominently hemosiderin-laden on SWI and thus labeled posthemorragic. Next, recent parenchymal lesions were defined as following: (i) edematous changes in the white matter or the cortex as visualized on FLAIR, foci of restricted diffusion on DWI; (ii) hemorrhagic lesions presenting as hyperintense on T1WI and blood-brain barrier breakdown translating as "blackblood" T1WI post-contrast enhancement. Finally, arterial evaluation searched for aneurysmal dilatations as well as disseminated or focal narrowings of the main branches of the circle of Willis on the TOF sequence. Arterial parietal enhancement was also assessed on "blackblood" T1WI postcontrast imaging, with caution regarding potential contamination of the high signal of adjacent veins (39). A suspicion of vasculitis was diagnosed on the basis of imaging if cerebral arteries demonstrated a stenosis or occlusion, and contrast enhancement of the vessel wall(40) .This suspicion was heightened if the abnormalities were associated with recent hemorrhagic, ischemic or edematous changes or contrast enhancement (40). In addition, the whole acquired volumes were screened for other abnormalities, including extra-axial masses and paranasal sinus pathology.

*2.2.2. Quantitative analyses of MRI data*

Cortical reconstruction and volumetric segmentation were performed with the Freesurfer 6 image analysis suite, which is documented and freely available for download online (http://surfer.nmr.mgh.harvard.edu/). The technical details of these procedures are described in prior publications, see (41-53). Briefly, this processing includes motion correction and averaging (Reuter et al. 2010) of volumetric T1WI, removal of non-brain tissue using a hybrid watershed/surface deformation procedure (51), automated Talairach transformation, segmentation of the subcortical white matter and deep gray matter volumetric structures (including hippocampus, amygdala, caudate, putamen, ventricles) (45, 46), intensity normalization (54), tessellation of the gray matter white matter boundary, automated topology correction (46, 55), and surface deformation following intensity gradients to optimally place the gray/white matters and gray matter/cerebrospinal fluid (CSF) borders at the location where the greatest shift in intensity defines the transition to the other tissue class (41-43). Once the cortical models are complete, a number of deformable procedures can be performed for further data processing and analysis including surface inflation (48), registration to a spherical atlas which is based on individual cortical folding patterns to match cortical geometry across subjects (47), parcellation of the cerebral cortex into units with respect to gyral and sulcal structure (46, 56), and creation of a variety of surface based data including maps of curvature and sulcal depth. This method uses both intensity and continuity information from the entire three-dimensional MR volume in segmentation and deformation procedures to produce representations of cortical thickness, calculated as the closest distance from the gray/white matters boundary to the gray matter/CSF boundary at each vertex on the tessellated surface (43). The maps are created using spatial intensity gradients across tissue classes and are therefore not simply reliant on absolute signal intensity. The maps produced are not restricted to the voxel resolution of the original data thus they can detect submillimeter differences between groups. Procedures for the measurement of cortical thickness have been validated against histological analysis (57)and manual measurements (58, 59). Freesurfer morphometric procedures have been demonstrated to show good test-retest reliability across scanner manufacturers and across field strengths (49, 53). Cortical volume and thickness maps obtained from Freesurfer recon-all pipeline were analyzed using mri_glmfit from the Freesurfer suite. Student t-tests were used to search for differences in maps of cortical volume and thickness between HIV+ subjects (taken individually or as a group) compared with HIV-PrEP users, and between HIV+ subjects and HIV-PrEP users (taken as separate or one group(s)) with that of healthy controls. The statistical maps were corrected for multiple comparison using the False Discovery Rate (FDR) as implemented in Freesurfer 6 and typically used for surface-based analyses, with the significance level set to $p<0.05$. We also selected a list of regions of interest (ROIs) (see Table 1) and extracted from Freesurfer's output their averaged volume and thickness. These data were then analyzed using JASP (Version 0.14.1) [Computer software]. All volumes were converted in percentage of total intracranial volume to account for global scaling of the ROI volumes with total brain volume. Student t-tests were used to search for differences in ROIs averaged volume (for cortical and subcortical ROIs) and thickness (for cortical ROIs only) between HIV+ subjects (taken individually or as a group) compared with HIV-PrEP users, and between HIV+ subjects and HIV-PrEP users (taken as separate or one group(s)) with that of healthy controls. Significance was considered at $p<0.05$ Bonferroni-corrected for the number of ROIs.

Table 1. Regions of Interest (ROIs)

| | |
|---|---|
| Cerebellum Cortex | Cuneus |
| Thalamus Proper | Enthorinal |
| Caudate | Fusiform |
| Putamen | Inferior parietal |
| Pallidum | Inferior temporal |
| Hippocampus | Isthmus cingulate |
| Amygdala | Lateral occipital |
| Accumbens area | Lateral orbitofrontal |
| Banks superior temporal sulcus | Lingual |
| Caudal anterior cingulate | Medial orbitofrontal |
| Caudal middle frontal | Middle temporal |

#### 2.3. FDG-PET data preprocessing and analyses

FDG-PET data were preprocessed and analyzed using the voxel-based Statistical Parametric Mapping software (SPM8, http://www. fil.ion.ucl.ac.uk/spm/, Wellcome Trust Centre for Neuroimaging, London, UK) based on conventional preprocessing, (individual and group level) subtractive and correlation analyses previously described (36, 60-64).

For that purpose, FDG-PET images were spatially normalized into the Montreal Neurologic Institute template (MNI, Montreal Neurologic Institute, Quebec, Canada) and then smoothed. Global activity normalization was performed by proportional scaling. For individual- and group-level subtractive analyses, we constructed general linear models (GLMs) of the preprocessed FDG-PET data of HIV+ subjects —taken individually or as a group—, and HIV-PrEP users or healthy controls taken as separate groups. Separate t-contrasts first identified brain areas where glucose metabolism was significantly lower or higher in HIV+ subjects (taken individually or as a group) compared with HIV-PrEP users. Then, similar group-level analyses compared the regional cerebral glucose metabolism of HIV+ subjects or HIV-PrEP users with that of healthy controls. One additional GLM also compared HIV+ subjects and HIV-PrEP users taken as one group with healthy controls. To search for a pathophysiological link between significant hypo- and hypermetabolic brain areas found in HIV+ subjects and HIV-PrEP users compared with healthy controls, we built GLMs comprising FDG-PET data of HIV+ subjects and HIV-PrEP users taken as one group, and the level of metabolism at significant hypometabolic voxels taken as covariates of interest.

To test the effects of RDU and TDF/FTC on regional cerebral glucose metabolism, we searched for voxels showing significant correlation with RDU score or TDF/FTC index using GLMs comprising FDG-PET data of HIV+ subjects and HIV-PrEP users taken as one group, and RDU score or TDF/FTC index taken as covariates of interest (separate analyses for RDU score and TDF/FTC index). Similar correlation analyses were also performed with the score of neuropsychological tests that significantly differed between HIV+ subjects or HIV-PrEP users. For significant voxels, regression plots and associated slope coefficients/p-values (Pearson's correlation) between glucose metabolism and covariates of interest were realized with Matlab (Version 9.2 R2017a, Mathworks Inc.; Natick, MA, USA). Results were considered significant at $p < .05$ corrected for multiple comparisons over the entire brain volume (Family Wise Error (FWE)). As the FWE rate may be considered too conservative at the individual level or for group-level analyses based on small samples sizes, results were also deemed significant at $p < .001$ uncorrected (height threshold: 0.001, cluster size $k \geq 50$ voxels) to minimize type II errors. This more liberal threshold was adopted only for voxels of brain areas previously reported as involved in the executive functions in healthy subjects or altered in HIV+ subjects (65-67). For voxel-based correlation analyses studies, results were also considered significant at small-volume-corrected $P<0.05$ using a 10-mm radius spherical volume of interest centered on the significant hypometabolic voxels.

### Supplementary Results

**Supplementary Table 2:** Brain areas showing significant decrease in metabolism in HIV+ subjects and HIV-PrEP users taken as one group

| | x | y | z | k | T | p |
|---|---|---|---|---|---|---|
| **R DLPFC** | 60 | 14 | 28 | 360 | 5.63 | 0.004 |
| **R DMPFC** | 16 | 64 | 4 | 2734 | 5.2 | 0.016 |
| | 30 | 58 | 12 | | 5.15 | 0.018 |
| | 46 | 48 | -6 | | 4.86 | 0.045 |
| **L PFC** | -6 | 66 | 6 | 582 | 4.49 | < 0.001 |
| | -4 | 62 | 22 | | 4.16 | < 0.001 |
| **L Insula** | -32 | 20 | -10 | 67 | 3.85 | < 0.001 |
| **L DLPFC** | -20 | 28 | 46 | 50 | 3.76 | < 0.001 |
| | -50 | 40 | 4 | 61 | 3.72 | < 0.001 |

R, right; L, left; DLPFC, dorso-lateral prefrontal cortex; DMPFC, dorso-medial prefrontal cortex; PFC, prefrontal cortex.

**Supplementary Table 3**

Brain areas showing significant increase of metabolism at group level (HIV+ and HIV-PrEP compared to HC)

| | x | y | z | k | T | p |
|---|---|---|---|---|---|---|
| **L Precuneus** | -12 | -50 | 72 | 1904 | 5.18 | 0.017 |
| **R Precuneus** | 12 | -64 | 64 | 1685 | 4.37 | <0.001 |
| **R primary somatosensory cortex** | 12 | -36 | 60 | 1685 | 5.04 | 0.026 |
| **R posterior cingulate cortex** | 16 | -26 | 44 | 1685 | 5.04 | < 0.001 |
| **R Occipital cortex** | 18 | -88 | 34 | 445 | 4.51 | < 0.001 |
| **L Occipital cortex** | -8 | -76 | 18 | 719 | 3.69 | < 0.001 |
| **L superior temporal gyrus** | -56 | -32 | 6 | 126 | 4.37 | < 0.001 |

R, right; L, left;

5. Slama H, Fery P, Verheulpen D, Vanzeveren N, Van Bogaert P. Cognitive Improvement of Attention and Inhibition in the Late Afternoon in Children With Attention-Deficit Hyperactivity

Disorder (ADHD) Treated With Osmotic-Release Oral System Methylphenidate. J Child Neurol. 2015;30(8):1000-9.

6. MacLeod CM. Half a century of research on the Stroop effect: an integrative review. Psychol Bull. 1991;109(2):163-203.

7. Vandierendonck A, Liefooghe B, Verbruggen F. Task Switching: Interplay of Reconfiguration and Interference Control. Psychological Bulletin. 2010;136(4):601-26.

8. Kiesel A, Wendt M, Jost K, Steinhauser M, Falkenstein M, Philipp AM, et al. Control and Interference in Task Switching-A Review. Psychological Bulletin. 2010;136(5):849-74.

9. Kreyszig E. Advanced engineering mathematics. 4th ed. New York: Wiley; 1979. xvii, 939, 92 p. p.

10. Baddeley AD, Hitch G. Working Memory. Psychology of Learning and Motivation. 1974;8:47-89.

11. Wechsler D. Manual for the Wechsler Intelligence Scale for Adults—Fourth edition (French norms). Paris, France: ECPA; 2011.

12. Kessels RP, van den Berg E, Ruis C, Brands AM. The backward span of the Corsi Block-Tapping Task and its association with the WAIS-III Digit Span. Assessment. 2008;15(4):426-34.

13. Woods DL, Kishiyamaa MM, Lund EW, Herron TJ, Edwards B, Poliva O, et al. Improving digit span assessment of short-term verbal memory. J Clin Exp Neuropsychol. 2011;33(1):101-11.

14. Corsi PM. Human memory and the medial temporal region of the brain: McGill University; 1972.

15. Fosco WD, Kofler MJ, Groves NB, Chan ESM, Raiker JS, Jr. Which 'Working' Components of Working Memory aren't Working in Youth with ADHD? J Abnorm Child Psychol. 2020;48(5):647-60.

16. Wager TD, Smith EE. Neuroimaging studies of working memory: a meta-analysis. Cogn Affect Behav Neurosci. 2003;3(4):255-74.

17. Zimmermann P, Fimm B. Tests d’Evaluation de l’Attention (TEA)(adaptation en français de Michel Leclercq). 2.2 ed: Psytest; 2006.

18. Buschke H, Fuld PA. Evaluating storage, retention, and retrieval in disordered memory and learning. Neurology. 1974;24(11):1019-25.

19. Rectem D, Poitrenaud J, Coyette F, Kalafat M, Van der Linden M. Une épreuve de rappel libre à 15 items avec remémoration sélective (RLS-15). In: Van der Linden M, GREMEM lmd, editors. L'évaluation des troubles de la mémoire : présentation de quatre tests de mémoire épisodique avec leur étalonnage. Marseille: Solal; 2004.

20. Baddeley A, Emslie H, Nimmo-Smith I. The Doors and People test: A test of visual and verbal recall and recognition. Bury St. Edmunds: Thames Valley Test Company; 1994.

26. Henguelle-Flamant C, Schaefer-Courcot F. "La tour de Londres": hypothèses théoriques et normalisation1994.

27. Cardebat D, Doyon B, Puel M, Goulet P, Joanette Y. [Formal and semantic lexical evocation in normal subjects. Performance and dynamics of production as a function of sex, age and educational level]. Acta Neurol Belg. 1990;90(4):207-17.

28. Bilocq V, De Partz MP, De Wilde V, Pillon A, Seron X, editors. Lexis - Test pour le diagnostic des troubles lexicaux chez le patient aphasique: De Boeck Solal; 2001.

29. Rey A. L'examen clinique en psychologie. Paris: Presses Universitaires de France; 1964.

30. Mahieux-Laurent F, Fabre C, Galbrun E, Dubrulle A, Moroni C, groupe de reflexion sur les praxies du CI-d-FS. [Validation of a brief screening scale evaluating praxic abilities for use in memory clinics. Evaluation in 419 controls, 127 mild cognitive impairment and 320 demented patients]. Rev Neurol (Paris). 2009;165(6-7):560-7.

31. McNair DM, Kahn RJ. Self-assessment of cognitive deficits. In: Crook T, Ferris S, Bartus R, editors. Assessment in Geriatric Psycho- pharmacology New Canaan (Conn.): Mark Powley; 1984.

32. Spielberger CD, Gorsuch RL, Lushene R, Vagg PR, Jacobs GA. Manual for the State-Trait Anxiety Inventory. Palo Alto, CA: Consulting Psychologists Press; 1983.
